# INFECTIVE ENDOCARDITIS AFTER ARRHYTHMIA DEVICE IMPLANTATION IN CARDIAC SURGERY PATIENTS

**DOI:** 10.1101/2025.11.25.25341028

**Authors:** Andreas Martinsson, Sigurdur Thorleifsson, Alice David, Araz Rawshani, Ulrika Snygg-Martin, Björn Redfors, Emma C Hansson, Amar Taha

## Abstract

**Background:** Patients undergoing cardiac surgery face an increased risk of infective endocarditis (IE), a risk that may be further exacerbated by implantation of cardiac implantable electronic devices (CIED). This study aimed to assess the risk of IE after CIED implantation within 30 days of cardiac surgery and identify associated risk factors.

**Methods:** All patients in Sweden, with no previous CIED, who underwent first-time cardiac surgery between 2006 and 2020 were included in this nationwide registry-based study. Cox proportional hazards models adjusted for demographic and clinical covariates were used to estimate the association between CIED implantation and the risk of IE.

**Results:** Out of 73,367 patients, 2,302 (3.1%) received a CIED within 30 days. Over a median follow-up of 6.5 years (interquartile range, 3.2–10.1 years), 1,556 patients (2.1%) developed IE. CIED implantation was associated with an increased IE risk (adjusted Hazard ratio (aHR) 1.46; 95% confidence interval (CI) 1.20-1.77). Patients with IE had a significantly higher risk of mortality compared to those without IE (aHR 1.80; 95% CI 1.67-1.94; p<0.001). The type of cardiac surgery influenced IE risk, with valvular surgeries showing a higher incidence than coronary artery bypass grafting. Risk factors independently associated with IE included male sex, age at surgery, diabetes mellitus, and peripheral vascular disease.

**Conclusions:** CIED implantation within 30 days after cardiac surgery is associated with an increased long-term risk of IE and subsequent mortality. The type of surgery and patient comorbidities influence the risk.

**CLINICAL PERSPECTIVE:** *What is new?:*

- In this large nationwide registry study, cardiac implantable electronic devices (CIEDs) placed within 30 days after cardiac surgery were associated with a substantial and persistent risk of infective endocarditis.
- The risk of infective endocarditis was influenced by the type of cardiac surgery and patient-specific factors, rather than the timing of device implantation.

*What are the clinical implications?:*

- Among post–cardiac surgery patients at elevated risk for infective endocarditis (e.g., those undergoing valvular procedures) who require a CIED, selecting devices associated with lower infection risk—such as leadless pacemakers or non-transvenous ICDs—may be a preferable strategy when appropriate.
- The optimal timing of CIED implantation after cardiac surgery remains uncertain. When delaying implantation, the rationale should center on allowing for potential recovery of conduction abnormalities, rather than attempting to mitigate the risk of device-related infection.

## INTRODUCTION

Arrhythmias necessitating cardiac implantable electronic device (CIED) implantation are relatively frequent among patients undergoing cardiac surgical procedures, with a notable rise in CIED implantation rates in this population over recent years.^1^ Pacemaker therapy constitutes the cornerstone of treatment for bradyarrhythmias.^2, 3^ While pacemakers significantly improve quality of life and survival^3^, their implantation is associated with a risk for cardiac implantable electronic device (CIED) infection, a potentially severe and life-threatening complication ^4, 5^ exerting a significant burden on healthcare resources.^6^

Infective endocarditis (IE), an infection of the endothelial surface of the heart, has shown a significant global rise in both the number of cases and associated mortality over the past three decades.^7^ Following CIED implant, the overall risk for CIED infection is approximately 1% during the first postoperative year.^8^ However, the risk of CIED-associated IE (CIED-IE) specifically in cardiac surgery patients is not well defined. Cardiac surgery patients, particularly those with prosthetic valves, are at an elevated risk for IE.^9, 10^ CIED implantation may further amplify this risk, as the device and its leads can serve as potential sites for infection, especially when introduced during the perioperative period. However, the exact risk of IE in patients receiving a CIED in the early postoperative period following cardiac surgery remains unclear. This study aims to address this gap by investigating the risk of CIED-IE in this specific patient population. Additionally, this study aimed to explore the risk factors associated with the development of IE in patients who are implanted with a CIED during the first 30 days following cardiac surgery.

## METHODS

### Study cohort

The study included all patients aged 18 years or older who underwent cardiac surgery in Sweden between January 2006 and December 2020. Eligible procedures were isolated coronary artery bypass grafting (CABG), isolated aortic valve surgery, isolated mitral valve surgery, isolated tricuspid valve surgery, or combinations of these procedures. Exclusion criteria were cardiac surgeries other than those mentioned above, a history of previous IE, CIED implantation before surgery, and death within the first 30 postoperative days. All patients had to survive at least 30 days after surgery, as the aim was to evaluate the risk associated with CIED implantation within this period. A flowchart of the included and excluded patients is depicted in Figure 1.

**Figure 1.**
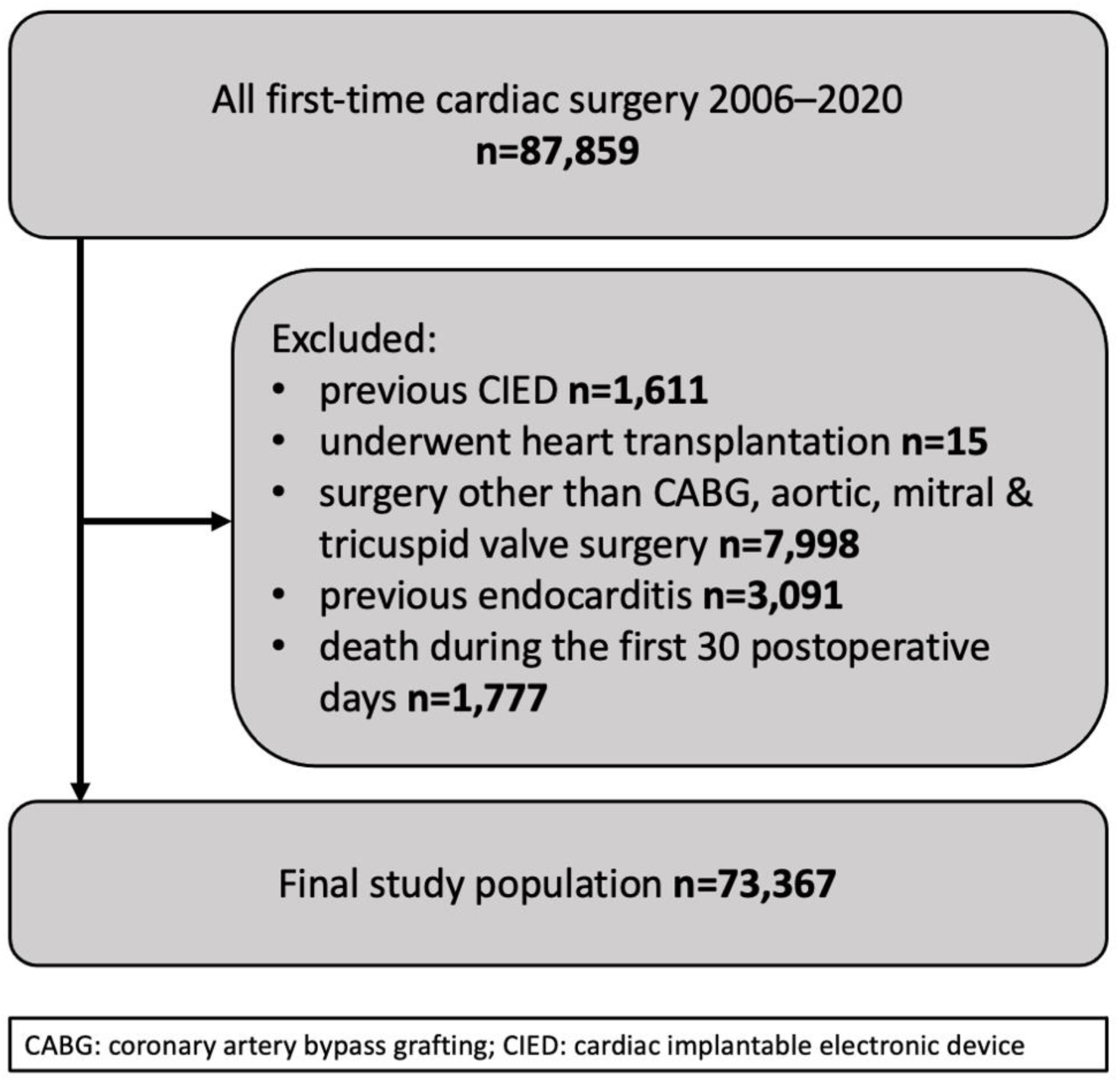
Flow chart of included and excluded patients

### Data sources

The data in the current study were derived from nationwide Swedish registries, covering all patients who underwent cardiac surgery in Sweden during the study period. The Swedish Cardiac Surgery Registry, a part of the Swedish Web-system for Enhancement and Development of Evidence-based care in Heart disease Evaluated According to Recommended Therapies (SWEDEHEART) registry, was used to identify the study population. The registry contains information on all cardiac surgeries performed in Sweden. It has been used since 1992, with excellent data validity and quality.^11, 12^ The Swedish ICD & Pacemaker Registry was used to collect implant data including implant date and type of CIED. The Swedish ICD & Pacemaker Registry collects data from all CIED implanting centers. Internal validation of the registry is continually monitored, and the register is also used as a clinical tool in daily practice.^13^ Comorbid conditions were collected from the mandatory National Patient Registry, which contains all diagnoses registered at any hospital in Sweden and forms the basis of financial reimbursement to departments and hospitals. All diagnoses during the study period were based on the International Classification of Diseases (ICD, 10th revision), the ICD-10 codes used can be found in Supplementary Table 1. The National Patient Registry has been validated with excellent coverage and validity.^14, 15^ Emigration and mortality data were collected from the Swedish Total Population Register which have complete coverage for deaths occurring within Sweden.^16^

In Sweden, all individuals receive a personal identification number; this number was anonymized and used to link all registries to specific individuals. The authors did not have access to individual personal identification numbers at any point. The manuscript was written according to the recommendation of the Strengthening the Reporting of Observational Studies in Epidemiology (STROBE) statement.^17^

### Outcomes

The primary objective was to evaluate the risk of IE within 30 days after surgery in patients with and without CIEDs. Furthermore, the study aimed to identify risk factors associated with an increased risk of IE. As a secondary objective, the study also evaluated the risk of endocarditis for patients with a CIED implanted at different time points before or after surgery to evaluate if the risk is particularly increased in temporal proximity to cardiac surgery. Additionally, other risk factors for long-term IE were evaluated.

### Statistics

Continuous variables were presented as means with standard deviation (SD) or medians with interquartile range (IQR) depending on their distribution. Categorical variables were presented as numbers and percentages. Incidence rates were calculated by dividing the number of events by follow-up years. 95% confidence intervals (95%CI) were calculated under the assumption of a Poisson distribution. Cumulative incidence functions were used to illustrate incidence of events over time to account for competing risks and achieve estimates of cause-specific risk. To calculate cause-specific hazard ratios adjusted Cox regressions models were used. These models allowed us to obtain hazard ratios (HR) with 95% CI. The Cox regression models were adjusted for factors assessed to influence both the risk of pacemaker implantation and the outcome. The following possible confounders were adjusted for: age at operation, sex, left ventricular ejection fraction, or a history of the following: myocardial infarction, diabetes, hypertension, peripheral artery disease, heart failure, chronic respiratory disease and atrial fibrillation. The adjustments were based on known clinical risk factors established from previous studies.^18^ The analysis were adjusted for multiple testing according to the Bonferroni-Holm method. There were only missing data for left ventricular ejection fraction, n=428 (0.6%) and body mass index n=5,022 (6.8%). Missing data was handled by multiple imputation by polytomous regression or logistic regression depending on the type of data. To assess collinearity variance inflation factor was used, there was no indication of collinearity in the models. Proportional hazards were tested using scaled Schoenfeld residuals and the model did meet the assumption of proportional hazards. Risk factors for subsequent endocarditis in patients with a pacemaker implanted were evaluated in a Cox regression model, first in a univariable fashion and thereafter in an adjusted model. As above, the risk factors selected for evaluation were based on results from previous studies and clinical experience.^17^ The outcome analyses started 30 days after surgery and any events prior were not excluded from the analyses. In a sensitivity analysis, the long-term risk of endocarditis associated with pacemaker implantation within one year was evaluated; in this analysis, pacemaker implantation was modelled using a time-updated approach. To compare the risk of CIED implantation within 30 days or 31-365 days after surgery, CIED implantation within these timeframes was separated, specifically for this analysis, patients had to survive the first 365 days.

All analyses were performed using R version 4.2.3 (R Foundation for Statistical Computing, Vienna, Austria), and all tests were two-tailed and interpreted at the 0.05 significance level.

### Ethical considerations and data availability statement

The study complied with the Declaration of Helsinki and was approved by the Swedish Ethical Review Authority (registration number 2021-00122, approved March 31, 2021). The data underlying this article will be available on reasonable request to the corresponding author, with permission of The Swedish Ethical Authority, The SWEDEHEART Registry and The Swedish Pacemaker/ICD Registry.

## RESULTS

### General

After exclusion criteria were applied, 73,367 patients were included in the study (Figure 1). The mean age (SD) was 68.0 (10.5) years, and 25.0% were women. In total, 3.1% (n=2,302) received a CIED within 30 days following cardiac surgery, 2.7% (n=1,957) received a pacemaker, and 0.4% (n=345) received an ICD. Patients who received a CIED were generally older, and more often had heart failure, atrial fibrillation and worse left ventricular ejection fraction (Table 1). Patients who had an ICD implanted had a higher prevalence of heart failure, low LVEF and previous myocardial infarction compared to the other groups. The variance inflation factor was below 2.0 for all included variables in the adjusted analyses.

**Table 1.** Baseline characteristics of patients with and without a CIED implanted within the first 30 postoperative days.

|  | No CIED<br>(N=71065) | ICD<br>(N=345) | Pacemaker<br>(N=1957) | Overall<br>(N=73367) |
| --- | --- | --- | --- | --- |
| **Sex** |  |  |  |  |
| Female | 17645 (24.8%) | 37 (10.7%) | 627 (32.0%) | 18309 (25.0%) |
| Male | 53420 (75.2%) | 308 (89.3%) | 1330 (68.0%) | 55058 (75.0%) |
| **Age** |  |  |  |  |
| Mean (SD) | 68.0 (10.4) | 66.1 (9.3) | 70.0 (11.2) | 68.0 (10.5) |
| **Body mass index** |  |  |  |  |
| Mean (SD) | 27.4 (6.2) | 27.9 (8.7) | 26.7 (4.7) | 27.4 (6.2) |
| Missing | 4847 (6.8%) | 31 (9.0%) | 144 (7.4%) | 5022 (6.8%) |
| **LVEF** |  |  |  |  |
| >50% | 50494 (71.1) | 77 (22.3) | 1324 (67.7) | 51895 (70.7) |
| <50% | 20158 (28.4%) | 267 (77.4%) | 619 (31.6%) | 21044 (28.7%) |
| Missing | 413 (0.6%) | 1 (0.3%) | 14 (0.7%) | 428 (0.6%) |
| **Heart failure** | 15328 (21.6%) | 220 (63.8%) | 655 (33.5%) | 16203 (22.1%) |
| **Atrial fibrillation** | 25788 (36.3%) | 142 (41.2%) | 1115 (57.0%) | 27045 (36.9%) |
| **Prior myocardial infarction** | 26279 (37.0%) | 220 (63.8%) | 386 (19.7%) | 26885 (36.6%) |
| **Chronic respiratory failure** | 7805 (11.0%) | 34 (9.9%) | 233 (11.9%) | 8072 (11.0%) |
| **Diabetes mellitus** | 17771 (25.0%) | 103 (29.9%) | 384 (19.6%) | 18258 (24.9%) |
| **Hypertension** | 46680 (65.7%) | 202 (58.6%) | 1218 (62.2%) | 48100 (65.6%) |
| **Peripheral vascular disease** | 9902 (13.9%) | 56 (16.2%) | 360 (18.4%) | 10318 (14.1%) |
| **Type of surgery** |  |  |  |  |
| Isolated CABG | 41674 (58.6%) | 254 (73.6%) | 304 (15.5%) | 42232 (57.6%) |
| Isolated aortic valve surgery | 15023 (21.1%) | 35 (10.1%) | 748 (38.2%) | 15806 (21.5%) |
| Isolated mitral valve surgery | 3651 (5.1%) | 16 (4.6%) | 200 (10.2%) | 3867 (5.3%) |
| Isolated tricuspid valve surgery | 200 (0.3%) | 0 (0%) | 15 (0.8%) | 215 (0.3%) |
| Combined valve and/or coronary surgery | 10517 (14.8%) | 40 (11.6%) | 690 (35.3%) | 11247 (15.3%) |
| Data are presented as mean (standard deviation, SD), or number (percentage)<br>CABG: coronary artery bypass grafting, CIED: cardiac implantable electronic device, ICD: implantable cardioverter-defibrillator,<br>LVEF: left ventricular ejection fraction |  |  |  |  |

### Risk of infective endocarditis in patients with CIED within 30 days

Over a median follow-up period of 6.4 years (IQR 3.2–10.1 years), 1,495 patients (1.9%) developed IE in the study population. A detailed description of patients with and without IE is shown in Supplementary Table S2. The cumulative incidence of IE in patients who received a CIED within the first 30 postoperative days was 1.0% (95% CI 0.7%-1.5%, Figure 2) at 1 year. The incidence of IE was highest during the first years after surgery, after which it remained stable during follow-up (Figure 2). While the incidence peaked early after surgery in patients with and without CIEDs, patients without a CIED had a more gradual decline than those with a CIED. Patients who had an ICD implanted had a similar cumulative incidence compared to those who had no device. Among patients who received a CIED within the first 30 days after surgery, those who developed incident IE more frequently had heart failure, atrial fibrillation, and peripheral vascular disease at baseline (Supplementary Table S2). Patients who received a CIED within 30 days showed a significantly higher risk of IE compared to those without a CIED (adjusted hazard ratio (aHR) 1.46, 95% CI 1.20-1.77; p<0.001). The association was stronger for ICDs (aHR 1.87, 95% CI 1.47-3.62, p=0.006) compared to conventional pacemakers (aHR 1.41, 95% CI 1.47-1.73, p=0.001)

**Figure 2.**
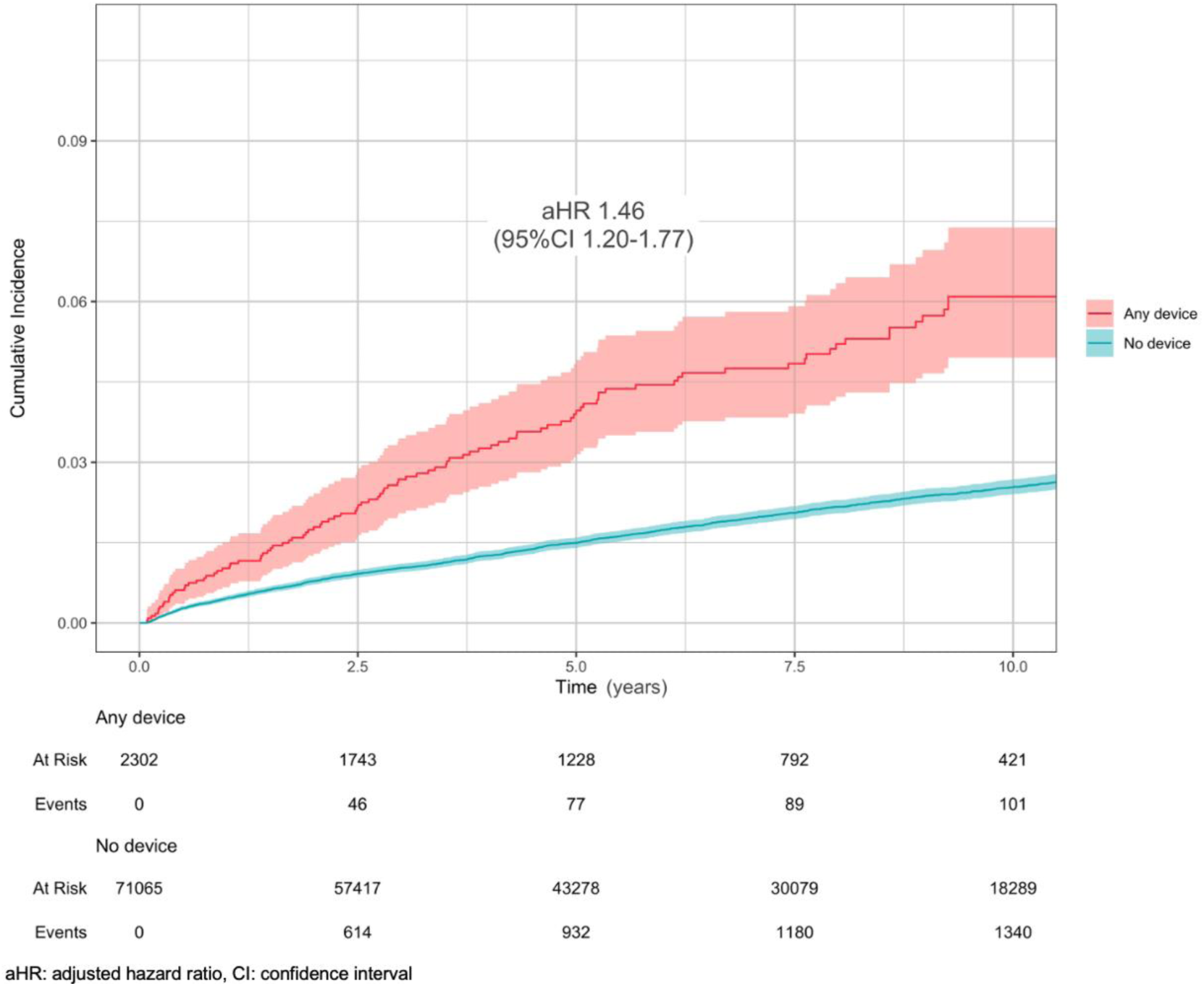
The cumulative incidence of infective endocarditis in patients with and without a pacemaker implanted within the first 30 postoperative days

There was no significant interaction between the risk for IE associated with CIED implantation and baseline variables, including age, sex, low ejection fraction, atrial fibrillation and diabetes mellitus, as shown in Supplementary Figure 1.

Developing IE during follow-up was associated with a significantly higher mortality compared to patients without IE (aHR 1.80, 95% CI 1.67-1.94; p<0.001) in a time-updated analysis. However, the risk of all-cause mortality after IE was higher for those without a CIED (aHR 1.87, 95% CI 1.73-2.01, p<0.001) than those with a CIED implanted within 30 days from the cardiac surgical procedure (aHR 1.06, 95% CI 0.77-1.47, p=0.72, p for interaction=0.001).

### Risk of IE and timing of CIED implantation

In a secondary analysis, the risk of IE and the timing of CIED implantation was evaluated by comparing placement within 30 days (n=2,079, HR 1.53, 95% CI 1.21-1.94, p<0.001) and between 31-365 days (n=884, HR 1.78, 95% CI 1.22-2.62, p=0.003). There was no significant difference in the risk of IE if a CIED was implanted during the first 30 postoperative days or within 31-365 days (p=0.49). The timing of implantation was also assessed for 0-7 days (n=966, HR 1.35, 95% CI 0.98-1.85, p=0.064) and >7 to 30 days (n=1,336, HR 1.55, 95% CI 1.18-2.02, p<0.001) with no statistical difference between the two groups (p=0.50). Additionally, patients with CIED placed prior to cardiac surgery (n=1,611) were compared to patients with CIED implanted 0-30 days after surgery; there was an association between previous CIED and risk of endocarditis (aHR 1.54, 95% CI 1.18-2.02, p=0.002), but there was no significant difference between the two groups (p=0.66).

### Type of surgery and IE

In patients who had a CIED implanted after cardiac surgery, the event rates and risk of IE differed substantially between different types of cardiac surgeries (Tables 2 and 3). Coronary surgery carried the least risk, while valvular procedures had higher event rates and adjusted risk estimates. To adjust for competing risk, cumulative incidence was evaluated. The results were similar as for the adjusted risk estimates, with the lowest incidence in coronary surgery patients, while aortic valve surgery and tricuspid valve surgery had higher cumulative incidence of IE (Figure 3). The cumulative incidence was 0.4% (0.1-1.2%) and 1.2% (0.4-2.61%) for isolated CABG at one and 10-year respectively, 1.2% (0.6-2.2%) and 8.9% (6.5-12.0%) for isolated aortic surgery, 1.0% (0.2-3.2%) and 6.8% (3.3-12.0%) for isolated mitral valve surgery, 0.0% (0.0-0.0%) and 14.0% (2.1-38.0%) for isolated tricuspid valve surgery, and 1.4% (0.7-2.5%) and 6.9% (4.9-9.4%) for combined cardiac surgery.

**Figure 3.**
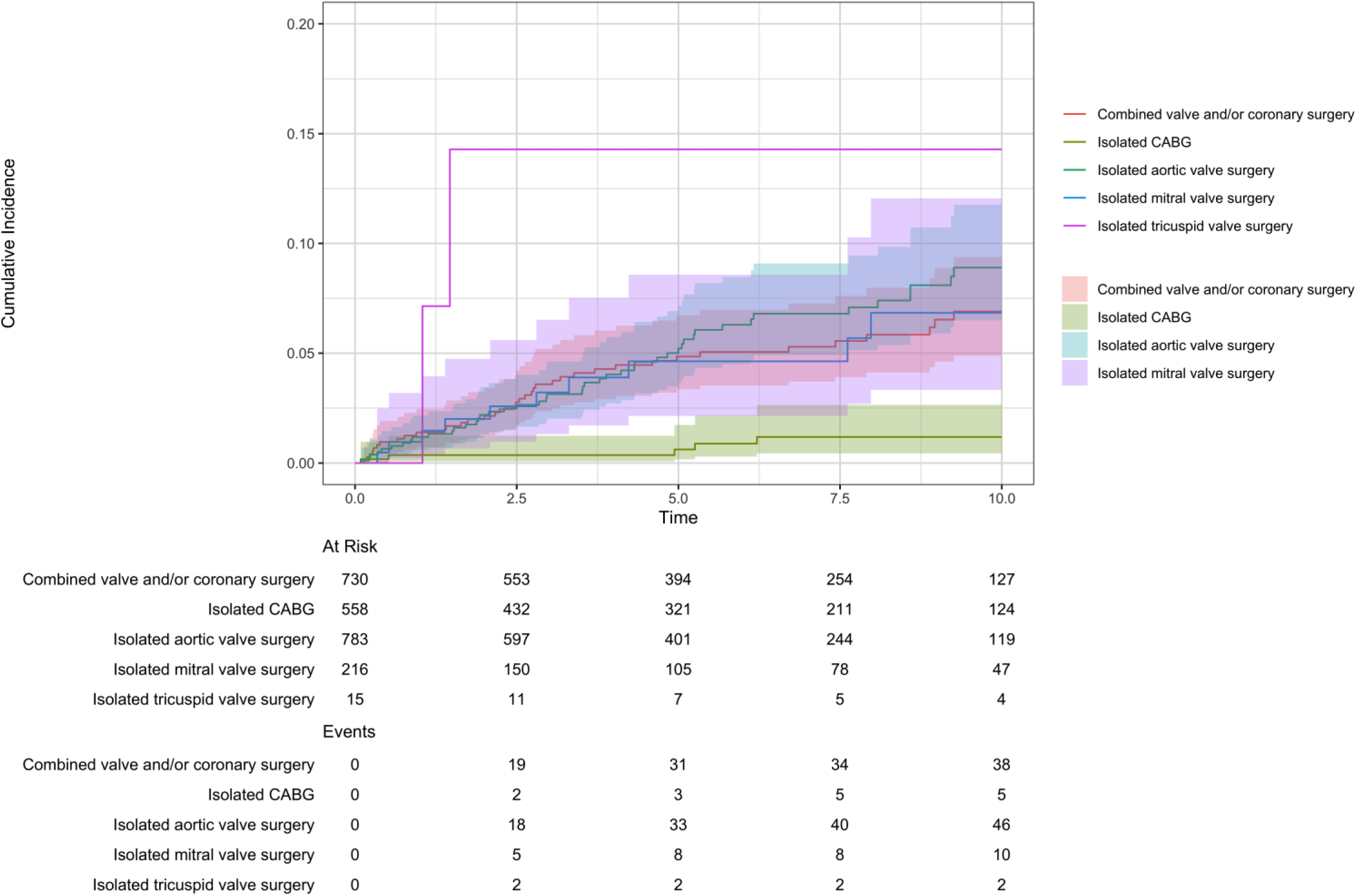
The cumulative incidence of infective endocarditis in patients who received a pacemaker within 30 postoperative days, according to the type of cardiac surgery

**Table 2.** Risk factors associated with infective endocarditis in patients who received a CIED within 30 days after cardiac surgery.

| Clinical characteristics | HR | 95% CI | p-value | Bonferroni-Holm<br>adjusted p-values |
| --- | --- | --- | --- | --- |
| Age (per year increase) | 1.01 | 1.01-1.02 | <0.001 | 0.001 |
| Sex (male) | 1.51 | 1.34-1.70 | <0.001 | <0.001 |
| Diabetes | 1.37 | 1.21-1.55 | <0.001 | <0.001 |
| Peripheral vascular disease | 1.26 | 1.11-1.43 | <0.001 | 0.005 |
| BMI (per SD increase) | 1.00 | 0.95-1.05 | 0.94 | 1.00 |
| Heart failure | 1.21 | 1.07-1.37 | 0.002 | 0.046 |
| Hypertension | 1.14 | 1.02-1.27 | 0.023 | 0.37 |
| Atrial fibrillation | 1.14 | 1.03-1.27 | 0.012 | 0.20 |
| Prior myocardial infarction | 1.01 | 0.87-1.17 | 0.90 | 1.00 |
| Chronic respiratory disease | 1.13 | 0.97-1.31 | 0.13 | 1.00 |
| Preoperative left ventricular<br>ejection fraction <50% | 0.94 | 0.83-1.06 | 0.33 | 1.00 |
| Type of surgery (CABG reference) |  |  |  |  |
| Aortic valve | 18.3 | 11.3-16.8 | <0.001 | <0.001 |
| Mitral valve | 9.8 | 7.5-12.8 | <0.001 | 0.045 |
| Tricuspid valve | 8.5 | 3.1-23.2 | <0.001 | 1.00 |
| Combined cardiac surgery | 13.8 | 11.3-16.8 | <0.001 | <0.001 |
| BMI: body mass index, CABG: coronary artery bypass grafting, CI: confidence interval; CIED: cardiac implantable electronic device HR: hazard ratio; SD: standard deviation |  |  |  |  |

**Table 3.** Event rate (with a 95% CI) of infective endocarditis per 100 patients-years in patients with and without a CIED according to the type of cardiac surgery.

| Type of surgery | Event rate per 100 patient-years<br>(Patients with CIED) | Event rate per 100 patient-years<br>(Patients without CIED) |
| --- | --- | --- |
| CABG | 0.15 (0.01-0.36) | 0.05 (0.04-0.06) |
| Aortic valve | 1.23 (0.91-1.63) | 0.89 (0.83-0.96) |
| Mitral valve | 1.05 (0.53-1.89) | 0.43 (0.35-0.53) |
| Tricuspid valve | 3.54 (0.43-12.80) | 0.21 (0.02-0.74) |
| Combined cardiac surgery | 1.00 (0.72-1.37) | 0.76 (0.69-0.83) |
| CABG: coronary artery bypass grafting, CI: confidence interval, CIED: cardiac implantable electronic device |  |  |

### Factors associated with IE in patients with CIED within 30 days

In the adjusted model, several additional factors were associated with an increased risk of IE (Table 2). The factors most strongly associated with IE were male sex (aHR 1.51, 95% CI 1.34-1.70) and diabetes mellitus (aHR 1.37, 95% CI 1.21-1.55), while age at surgery (aHR 1.01, 95% CI 1.01-1.02) per year increase), peripheral vascular disease (aHR 1.26, 95% CI 1.11-1.43), heart failure (aHR 1.21, 95% CI 1.07-1.37), hypertension (aHR 1.14, 95% CI 1.02-1.27), and atrial fibrillation (aHR 1.14, 95% CI 1.03-1.27) had a more moderate association. When applying the Bonferroni method to adjust for multiple testing, the association between IE and atrial fibrillation and hypertension was no longer statistically significant.

## DISCUSSION

The most significant findings in this study are that the placement of a CIED increases the long-term risk of IE after cardiac surgery, and that the timing of device placement was not associated with an increased long-term risk of IE. Both defibrillators and conventional pacemakers increased the risk of subsequent IE; no statistical difference was found between the two types of CIEDs. In addition, the type of surgery performed had a strong effect on the accompanying risk of IE, both in patients with and without CIED placement.

The current study reports on contemporary outcomes associated with CIED placement at the time of cardiac surgery and subsequent risk of IE. The risk of CIED placement is non-negligible after cardiac surgery but varies greatly with the type of procedure.^1^ Consistent with previous reports, the long-term risk of IE in this study also varied with surgical procedure, being more common after valvular surgery.^19^ Several mechanisms may explain how CIED placement contributes to increased endocarditis risk, including venous instrumentation during the intervention and the permanent presence of foreign material in close contact with cardiac tissue. Together, these factors may account for both the early divergence and the sustained excess incidence of IE observed in this study.

Although the occurrence of bradyarrhythmias often necessitates pacemaker implantation, these findings highlight the need for careful evaluation of device indications in high-risk patient populations. In patients at elevated risk of postoperative bradyarrhythmias (e.g., multiple valve surgery and pre-existing conduction abnormalities), placement of epicardial pacing leads during the index procedure may reduce subsequent IE risk. ^20^ An alternative pacing strategy is the use of leadless pacemakers, which can provide both atrial and ventricular pacing.^21–23^ Although randomized trial data are lacking, real-world evidence suggests that leadless pacemakers carry a very low risk of device-related infections and may represent a valuable option for post-cardiac surgery patients.^24^. Similarly, in post-cardiac surgery patients with a risk of IE (e.g. valvular surgery) and a need for an ICD, the use of non-transvenous ICDs (e.g. the subcutaneous ICD) might be a preferable option when pacing is not required.^25^

For those who had a CIED implanted, the associated risk did not seem to differ depending on patient characteristics, indicating that the type of surgery and the additional intervention and material in the venous system may have played a more central role. However, it should be noted that the baseline risk differs depending on comorbidities and patient characteristics, which have previously been shown to increase the risk of IE after CIED implantation.^17^ In the current study, male sex and diabetes had the most prominent association with increased risk of IE.

In accordance with previous studies, a diagnosis of IE results in poor long-term outcomes and a clear association with increased risk of death.^3–4^ This finding is unsurprising but should be emphasized as IE is a condition with substantial mortality and morbidity. Consistent with the findings of a previous large study, all-cause mortality following IE was higher among patients without a CIED than among those with one.^26^ This may be explained by the fact that, in CIED-associated IE, the infection typically involves the right side of the heart, which carries a lower risk of arterial embolization. In addition, CIED-IE is often managed with lead extraction, a procedure associated with a significant reduction in mortality risk.^27^ In the current study, patients with endocarditis before or at the time of surgery were excluded; these could potentially represent a cohort with an even higher risk not only for IE but even a high risk for conduction system disease following cardiac surgery.

There is a great debate regarding the optimal timing of device placement after cardiac surgery. The current guidelines are based on only a handful, rather small reports, and current practice differs across healthcare providers.^2,28^ Cardiac surgery results in trauma and subsequent inflammation of the myocardial tissue, occasionally causing transient bradyarrhythmias which might potentially regress during the hospital stay. There has been speculation that CIED placement close to the time of surgery increases the risk of infection and might not allow adequate time for recovery of the conduction system after surgery. Although our study did not investigate the optimal timing of CIED implantation after cardiac surgery or whether a longer waiting period could reduce the need for a CIED, the timing of implantation did not appear to have a major impact on the risk of IE. The associated risk increase was similar between patients who received the device within 0–7 days and those implanted between 8–30 days postoperatively. Likewise, there was no clear difference between patients who had their device 30-365 days after surgery, nor prior to cardiac surgery. This suggests that, if adequate time has been given to allow residual surgical trauma and inflammation to subside, the risk of IE is not an important factor when deciding on the optimal timing of device implant. However, when combined, CIED presence and cardiac surgery seem to markedly increase the risk of IE.

### Strengths and limitations

The current study has important limitations that should be considered when evaluating the results, most importantly the retrospective design, which makes residual confounding a possibility and causal association cannot be ascertained. In addition, the first 30 days were excluded, and thus, patients who develop very early endocarditis are not considered. In contrast, the study also has central qualities, including the contemporary setting and the use of nationwide registries capturing >99% of all cardiac surgery and CIED implant procedures.

## Conclusion

The placement of a cardiac implantable electronic device after cardiac surgery is associated with an increased risk of infective endocarditis. The associated risk increase is similar irrespective of the type of surgery, but the baseline risk differs greatly depending on whether valvular surgery is performed or not. The timing of CIED placement did not influence the association.

## Data Availability

The data underlying this article will be available on reasonable request to the corresponding author, with permission of The Swedish Ethical Authority, The SWEDEHEART Registry and The Swedish Pacemaker/ICD Registry.

## Non-standard Abbreviations and Acronyms

aHR: adjusted hazard ratio
CABG: coronary artery bypass grafting
CIED: cardiac implantable electronic device
ICD: implantable-cardioverter defibrillator,
IE: infective endocarditis

